# PAGEANT: Personal Access to Genome and Analysis of Natural Traits

**DOI:** 10.1101/2021.10.11.21264790

**Authors:** Jie Huang, Zhi-Sheng Liang, Stefano Pallotti, Janice M. Ranson, David J. Llewellyn, Zhi-Jie Zheng, Dan A. King, Qiang Zhou, Houfeng Zheng, Valerio Napolioni

## Abstract

GWASs have identified numerous genetic variants associated with a wide variety of diseases, yet despite the wide availability of genetic testing the insights that would enhance the interpretability of these results are not widely available to members of the public. As a proof of concept and demonstration of technological feasibility, we developed PAGEANT (<u>P</u>ersonal <u>A</u>ccess to <u>Ge</u>nome & <u>A</u>nalysis of <u>N</u>atural <u>T</u>raits), usable through Graphical User Interface or command line-based version, aiming to serve as a protocol and prototype that guides the overarching design of genetic reporting tools. PAGEANT is structured across five core modules, summarized by five Qs: (1) Quality assurance of the genetic data; (2) Qualitative assessment of genetic characteristics; (3) Quantitative assessment of health risk susceptibility based on polygenic risk scores and population reference; (4) Query of third-party variant databases (e.g., ClinVAR and PharmGKB); and (5) Quick Response code of genetic variants of interest. Literature review was conducted to compare PAGEANT with academic and industry tools. For 2,504 genomes made publicly available through the 1,000 Genomes Project, we derived their genomic characteristics for a suite of qualitative and quantitative traits. One exemplary trait is susceptibility to COVID-19, based on the most up-to-date scientific findings reported.

## Introduction

The start of the millennium was marked by a significant achievement of human health research - the completion of the draft Human Genome Project. Over the past two decades, millions of human genomes have been sequenced and even many more have been genotyped. Starting from a single project to numerous publications, human genetic research is now serving more and more for patients and the general public.

Academia has thus far been the driving force for discovering genetic loci associated with complex traits, delivering thousands of genome-wide association studies (GWASs), and reporting millions of genetic loci plausibly associated with various diseases and health conditions. GWASs have grown from hundreds of participants to over a million [**1**], spanning a wide range of health phenotypes. Polygenic Risk Scores (PRS) based on GWAS results can incorporate millions of genetic variants to accurately predict individual risk of health conditions, with some offering superior predictive performance compared to established risk factors [**2,3**]. The concept of ‘big data’ for health is finally becoming actionable, in that genetic variation may have diagnostic or therapeutic implications.

A comprehensive review of the literature on well-studied common diseases/traits where PRS showed clinical value was recently conducted by Lewis and Vassos [**4**]. The predictive accuracy of the PRS has already demonstrated for common diseases including type 2 diabetes [**5**] and coronary heart disease [**6**]. Also, using data from the large-scale UK Biobank study [**7**], researchers from the United States, United Kingdom [**8**], and Pacific Islands [**9**] have generated GWAS results for thousands of traits and billions of data points; these findings provide new insights into disease risk [**10**]. However, the public lack the means to avail such data for interpretation of their own genomes. There is therefore a need for the design and development of user-friendly systems for delivering personalized genomic information, both for disease treatment and prevention, considering individual genetic variation, lifestyle, and environmental characteristics [**11**].

Several direct-to-consumer (DTC) companies offer genetic testing and reporting over the counter and online, with millions of users now having had their DNA assayed and received genetic reports [**12**]. Genetic testing is a key area for US government regulation agencies. The US National Institute of Health (NIH) National Center for Biotechnology Information (NCBI) Genetic Testing Registry contains over 60,000 genetic tests for over 10,000 conditions [**13**]. The US Food and Drug Administration has over 400 entries for pharmacogenomic biomarkers used in drug labeling and published a list of Direct-To-Consumer (DTC) tests with marketing authorization (https://www.fda.gov/medical-devices/in-vitro-diagnostics/direct-consumer-tests#list). Nevertheless, DTC genetic testing is under strict government regulation, and several important ethical concerns remain [**14**]. Concerns include possible psychological harm [**15**], lack of professional genetic counselling [**16**], lack of data protection [**17**] and lack of validity and clinical utility of test results [**18**].

Under the guidance of ethical principles especially related to genetic data confidentiality, we developed PAGEANT (<u>P</u>ersonal <u>A</u>ccess to <u>Ge</u>nome & <u>A</u>nalysis of <u>N</u>atural <u>T</u>raits), a self-completion genetic reporting tool for individuals with personal genomic data. PAGEANT follows five core philosophies: (1) Academic quality and standards. State-of-the-art algorithms incorporate millions of genetic variants to calculate individual Polygenic Risk Score (PRS); (2) Confidential data run locally, without the need to send genomic data to cloud servers; (3) Generalizable architecture and algorithm, where our “five-Q” design could easily grow from the basic version for dozens of traits to hundreds and thousands of traits; (4) Transparent source code for all underlying programming scripts; (5) User-centric, as users have full control to add or remove certain traits into or from a genetic report.

PAGEANT aims to provide proof of concept for a scientifically driven architecture with a user-friendly interface, offering a technologically feasible approach to allow users to understand their genetic traits and predictive value of an individual’s genomic variation.

## Methods

### Review and comparison with existing tools

With the aim of contextualizing PAGEANT in the present setting, we performed an extensive literature search on both PubMed and Google using the keywords: ‘‘personalized genome”, ‘‘third-party interpretation”, ‘‘genome interpretation”, ‘‘genome”, ‘‘genetic testing” and ‘‘risk prediction” applying the following algorithm: (genome interpretation OR genome) AND (third-party interpretation) AND (genetic testing OR risk prediction). The identification of eligible studies was not restricted to English language. Studies references were also analyzed to find any study not available from the electronic databases. We also determined whether each of the identified tools are still functional and available on the web until July 2021.

### Overarching design of the user interface

PAGEANT is an open-source, customizable platform with a version suitable for non-technical users. The basic version of PAGEANT has five modules, summarized by five Qs, described below.

1. *Quality control report of genetic data*. To our knowledge, PAGEANT is the only genetic reporting tool that first reports genotype quality before reporting genotype-derived results. This step is especially important for DTC users, to ensure quality control. PAGEANT generates a genotype quality control (QC) report for the input personal genome and also for thousands of genomes used in the reference panel later used for calculation of the PRS. PAGEANT takes as input a variant call format (VCF) file, generated using a variety of genotyping platforms, such as whole-genome sequencing or Single Nucleotide Polymorphism (SNP) array.
2. *Qualitative assessment of genetic characteristics of absolute or relatively high certainty*. We broadly divided the traits into qualitative and quantitative traits. Qualitative traits are categorical such as YES/NO or Presence/Absence or categorical such as blood type {A, B, AB, O}. The determination of qualitative traits is straightforward with a definitive outcome obtained from the presence of target variants. Examples include tagging a particular functional haplotype of broad clinical relevance (such as ABO blood type, *APOE* genotype, *FTO* flagship SNP rs9939609, etc.) or PLINK [**19**] metrics such as chromosome-level of heterozygosity and genetically derived sex.
3. *Quantitative assessment of health risk susceptibility based on PRS*. PAGEANT is pre-installed with a small number of complex traits that have high disease burden and strong evidence of genetic risk prediction. We used GWAS summary results from UK Biobank (UKB, http://www.nealelab.is/uk-biobank) and BioBank Japan (BBJ, http://jenger.riken.jp/en/result). Pruning methods were used to retain independent genetic variants (r^2^<0.1) and construct a scoring file for each trait. To allow users to interpret their own PRS in the context of a large population, PAGEANT uses the specified list of SNPs and statistical models to calculate PRS for the provided population reference, in addition to calculating PRS for individuals. The 1000 Genomes Project (G1K) genetic data (https://www.internationalgenome.org/data/)[**20**] are the foundation for most GWAS and PRS studies, and this is used as the default reference panel. PAGEANT defines three risk categories based on the position of the personal genome across the PRS distribution (<25% Low risk, 25-75% Normal, >75% high risk).
4. *Query of third-party variants databases such as ClinVAR [****21****] and PharmGKB [****22***]. This aims to increase PAGEANT’s generalizability. With increasing interest in precision health and guidelines for drug usage, SNPs that predict clinical pathogenicity and pharmacogenomic relevance are increasingly incorporated into genotyping array panels. At the same time, SNPs with detailed annotations are added into such databases constantly. This module establishes technical standards and facilitates a diverse range of genetic interpretation tools.
5. *Quick Response (QR) code generation for tagging individual genomes, guaranteeing personal privacy and quick retrieval of the PAGEANT genetic report*. Our group previously developed a SNP panel and an online tool for checking genotype concordance through comparing QR codes [**23**]. In that work we identified 80 “fingerprinting” SNPs that could be used to uniquely identify a person. We subsequently implemented a web-based tool to convert the genotype of those 80 SNPs into a QR code, so that users could use that QR code as a genetic ID for quick concordance check. Here, the QR code module in PAGEANT uses the same 80 SNPs as an example, to illustrate how a user could conveniently scan a list of SNPs coded and encrypted in a QR code to extract his personal genomic data for downstream usage.

### Technical implementation

The tool is written using Python v3; the *Pandas* module was used to read, clean, and analyze various data. The *Matplotlib* module was used for plotting. The *PyQt5* module was used for API related functionality (along with specific classes linked to Qt C++, it facilitates further graphical applications). The *Jinja2* module was used to generate the HTML report. Finally, we used the *Pyinstaller* module to organize core scripts and all dependencies into a single executable file, without the need to construct the running environment.

We also embedded two widely used bioinformatics tools. First, we used PLINK [**19**] to convert and filter the genotype data and process quality control. Second, we used VEP [**24**] to generate the various annotations for the input genotype data. In our default version, we also embedded two widely used genetic databases: ClinVar [**21**] and PharmGKB [**22**].

Finally, for the 5^th^ Q (QR code), we used existing python packages “qrcode” and “pyzbar” for encoding/decoding and “rsa” and “pyDes” for encryption/decryption. The encryption/decryption is based on an asymmetric cryptography algorithm.

Application Programming Interfaces (APIs) for projecting personal genome on population reference genomes and for generating QR code, based on a SNP list and public key, were written from scratch with commonly used python libraries. An API for adding rsID was modeled on a similar python script of Pheweb [**25**], with the added flexibility to specify the REF vs. ALT alleles. This API will foreseeably be replaced by standard genomic tools (i.e., bcftools) once the VCF format is widely used for GWAS [**26**]. The source code and example command line usage for all three APIs are presented on PAGEANT GitHub page (https://github.com/jielab/pageant).

## Results

### Review and comparison with existing tools

Through the literature search we identified a structured content analysis of 23 third-party interpretation tools conducted by Nelson and Fullerton published in 2018 [**27**], on which we decided to build our review and comparative analysis. Thus, we searched for tools that were made available to the public since the end of their review period (December 2016), identifying five additional third-party interpretation tools, namely Allelica, CodeGenEU, GenePlaza, Impute.me and Self Decode (**Table 1**).

**Table 1.** DTC genetic testing interpretation tools. For each identified tool we reported the developer type, country, website link and their status (active vs. deactivated) in July 2021. NA=Not Available.

| Name | Developer type | Country | Web-site link | Status |
| --- | --- | --- | --- | --- |
| Impute.me | Academic | Denmark | https://www.impute.me/ | Active |
| Infinome | Academic | USA | https://www.infino.me/ | Active |
| Interpretome | Academic | USA | NA | Deactivated |
| openSNP | Academic | Germany | https://opensnp.org/ | Active |
| Allelica | Company | Italy | https://www.allelica.com/ | Active |
| Anabolic Genes | Company | France | http://www.anabolicgenes.com/ | Deactivated |
| Athletigen | Company | Canada | https://athletigen.com/ | Active |
| CodeGenEU | Company | Europe (Not Specified) | https://codegen.eu/ | Active |
| DNA Doctor | Company | USA | http://www.biostatushealth.com/dnadoctor/ | Active |
| DNA Tribes | Company | USA | https://dnatribes.com/ | Active |
| DNA.land | Company | USA | https://dna.land/ | Active |
| DNAFit | Company | UK | https://www.dnafit.com | Active |
| Enlis Genome Personal | Company | USA | https://www.enlis.com/personal_edition.html | Active |
| Family Tree DNA | Company | USA | https://www.familytreedna.com | Active |
| GEDMatch | Company | USA | https://www.gedmatch.com/ | Active |
| GenePlaza | Company | France | https://www.geneplaza.com/ | Active |
| Genetic Genie | Company | USA | https://geneticgenie.org/ | Active |
| GENETICconcept/Oh My Genes | Company | France | http://fr.geneticconcept.com/index.html | Deactivated |
| Golden Helix Genome Browser | Company | USA | https://www.goldenhelix.com/products/GenomeBrowse/ | Active |
| GPS Origins | Company | UK | https://www.ibdna.com/tests/gps-origins/ | Active |
| Livewello | Company | USA | https://livewello.com/ | Active |
| NutraHacker | Company | USA | https://www.nutrahacker.com/ | Active |
| Promethase | Company | Israel | https://promethease.com/ | Active |
| Self Decode | Company | UK | https://selfdecode.com/ | Active |
| WeGene | Company | China | https://www.wegene.com/en/ | Active |
| David Pike's utilities | Non-specialist | Canada | https://www.math.mun.ca/~dapike/FF23utils/ | Active |
| GeneKnot | Non-specialist | NA | NA | Deactivated |
| James Lick Haplogroup Analysis | Non-specialist | NA | https://dna.jameslick.com/mthap/ | Active |

Among all the 28 tools that we reviewed, four (Interpretome, Anabolic Genes, GENETIConcept and GeneKnot) were deactivated, with AnabolicGenes and GENETIConcept being incorporated into a new company, named “Oh My Genes”, which appears inactive (**Table 1**). Since the main aim of PAGEANT is to provide an open-source, customizable platform for determining individual genetic-based risk profiles, based on reliable and transparent resource provided by the academic field, we focused our attention on tools available free of charge by academic-based providers.

Three tools categorised as academic-based providers by Nelson and Fullerton [**27**] were not considered as such in the current review. Promethase, a tool developed by the SNPedia team, requires a fee to pay (minimum $12) according to the number of reports requested by the user. Likewise, DNA.land [**28**], recently transitioned from an academic research project to a for-profit company, and the source code is not publicly available. Infino.me requires health (weight and blood pressure) and physical activity measures obtained from personally wearable tracker devices (e.g., FitBit, Withings) to get access to their genetic report.

Thus, we only considered Impute.me and openSNP for comparison with PAGEANT. Impute.me and openSNP were released in 2015 and in 2011, respectively, supported by companion papers, published in *PLoS One* in 2014 (for openSNP [**29**]) and in *Frontiers in Genetics* in 2020 (for Impute.me [**30**]). The main features of Impute.me and openSNP, compared to the ones offered by PAGEANT are reported in **Table 2**. These two tools provide publicly available source codes and easy-to-access websites. However, they have two main disadvantages: lack of customizability and data confidentiality concerns. Users are not be able to easily customize those tools including the number of traits and the number of SNPs for each trait. In contrast, PAGEANT allows users to customize the genetic report (such as adding/removing traits to be reported, change reference genome to be used) by uploading new files or creating new template-based folders. With regards to data confidentiality, openSNP consists of an open forum for public discussion about the results coming from the interpretation of individual SNPs, previously found to be associated with a certain trait in any of the GWAS carried out so far. The individual genetic data uploaded in openSNP is retained with the aim of providing a public discussion on the obtained results. This may raise important concerns regarding potentially misleading scientific communication within a lay audience.

**Table 2.**
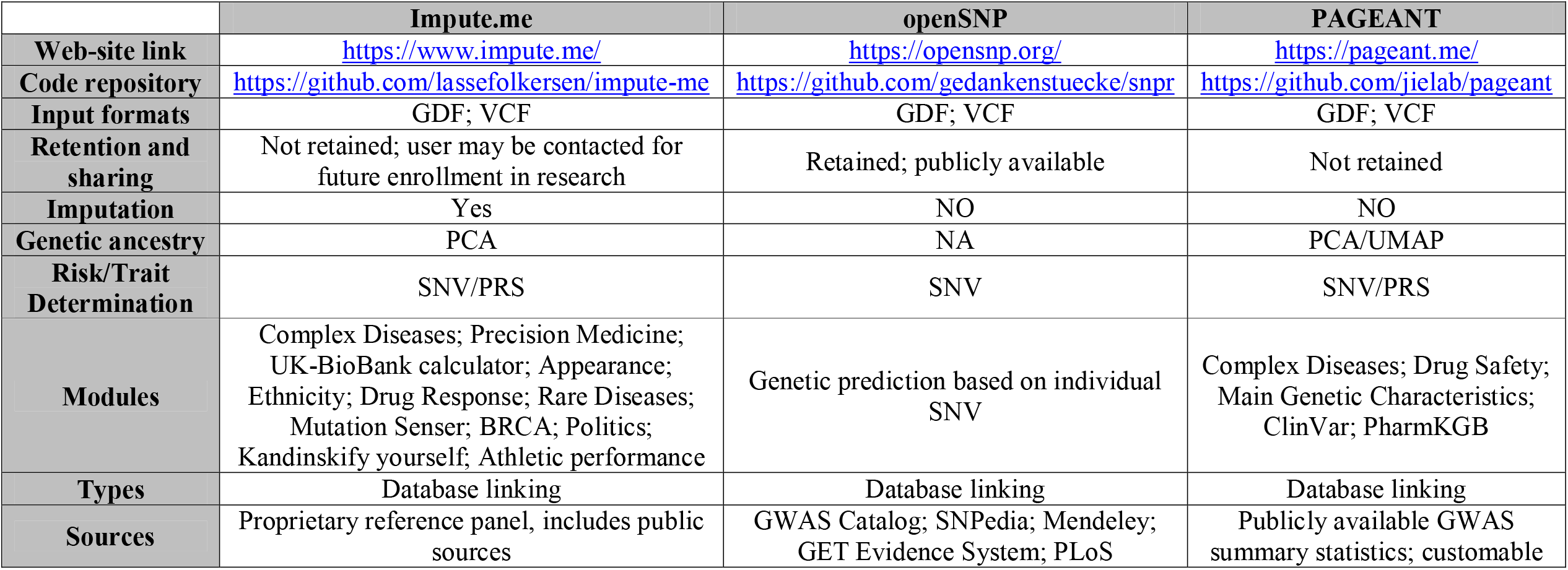
Comparison of PAGEANT with other “Academic” DTC genetic testing tools. GDF=genetic data file; NA=Not Available; PCA=Principal Component Analysis; PRS=Polygenic Risk Score; SNV=Single Nucleotide Variant; UMAP=Uniform Manifold Approximation and Projection.

Impute.me was the most similar resource when comparing it to PAGEANT. Thus, we decided to benchmark PAGEANT exclusively by comparing its performance with Impute.me. Compared to Impute.me, PAGEANT has the advantage of being a standalone tool that could be run on a laptop and without internet connection. In general, a website is more prone to security breach [**31**]. As its name implies, Impute.me actually impute users’ genetic data that that process takes up to several days on a personal computer with typical settings. With the sharp decrease of sequencing cost, imputation is likely to become obsolete in the near future. For example, the UK Biobank project is scheduled to conduct whole genome sequencing for all 500,000 participants.

### The “5-Q” modules of PAGEANT

The technical implementation of the five-Q modules is shown in **Figure 1**. PAGEANT is a suite of common bioinformatics software including PLINK [**19**] and VEP [**24**] to manage and annotate user provided genetic data. The main python script is used to generate the user interface, manage the process and data flow, and eventually generate an easy-to-read report. **Figure 2** outlines the file structures when the software is locally installed. Advanced users could work on the folders directly to customize some of the underlying databases and the scope of traits to be reported. The graphical user interface (GUI) was designed in such a way that users could fully customize various parameters before running the full program. It allows users to obtain an example genetic report after loading the GUI interface, by clicking the “Analyze” button at the bottom of the “I/O” page (**Figure 3A**), after selecting the “Reference population ethnical group” in the “Quantitative” page (**Figure 3B**), a necessary step to obtain reliable PRS scores. The GUI page for the five-Q modules is preloaded with default links to key directories and software parameters, which can be customized by advanced users. Advanced users can also customize their genetic report by (1) editing the configuration file; (2) adding/removing traits to be tested, and (3) replacing PRS reference scoring files. To enhance the security and the confidentiality of data processing, even if PAGEANT does not store user’s data, we implemented three APIs that allow users to access three key components of PAGEANT (**Figure 3C**). These three APIs could be run standalone, either through the GUI interface or through the command line.

**Figure 1.**
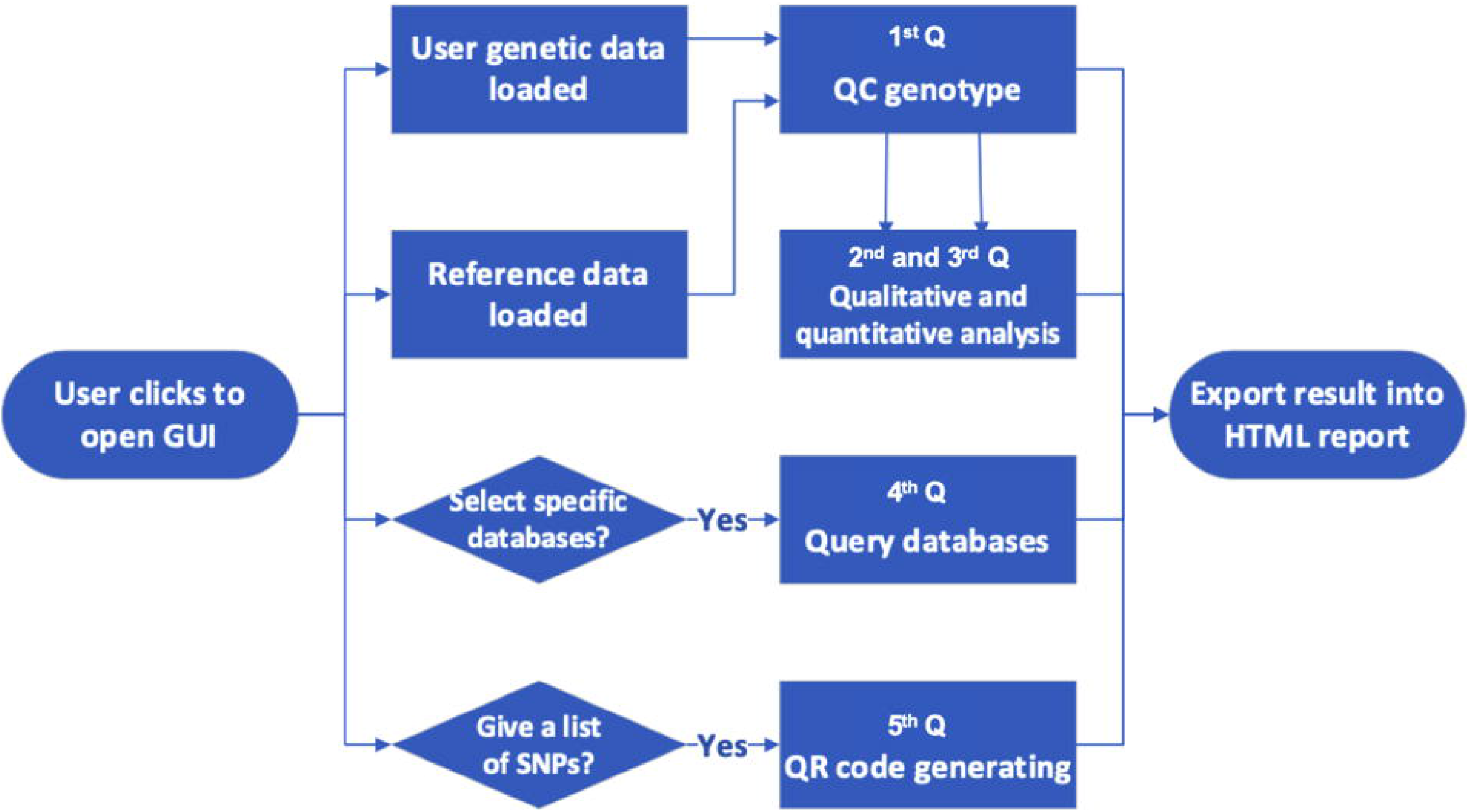
The technical implementation of the five-Q modules.

**Figure 2.**
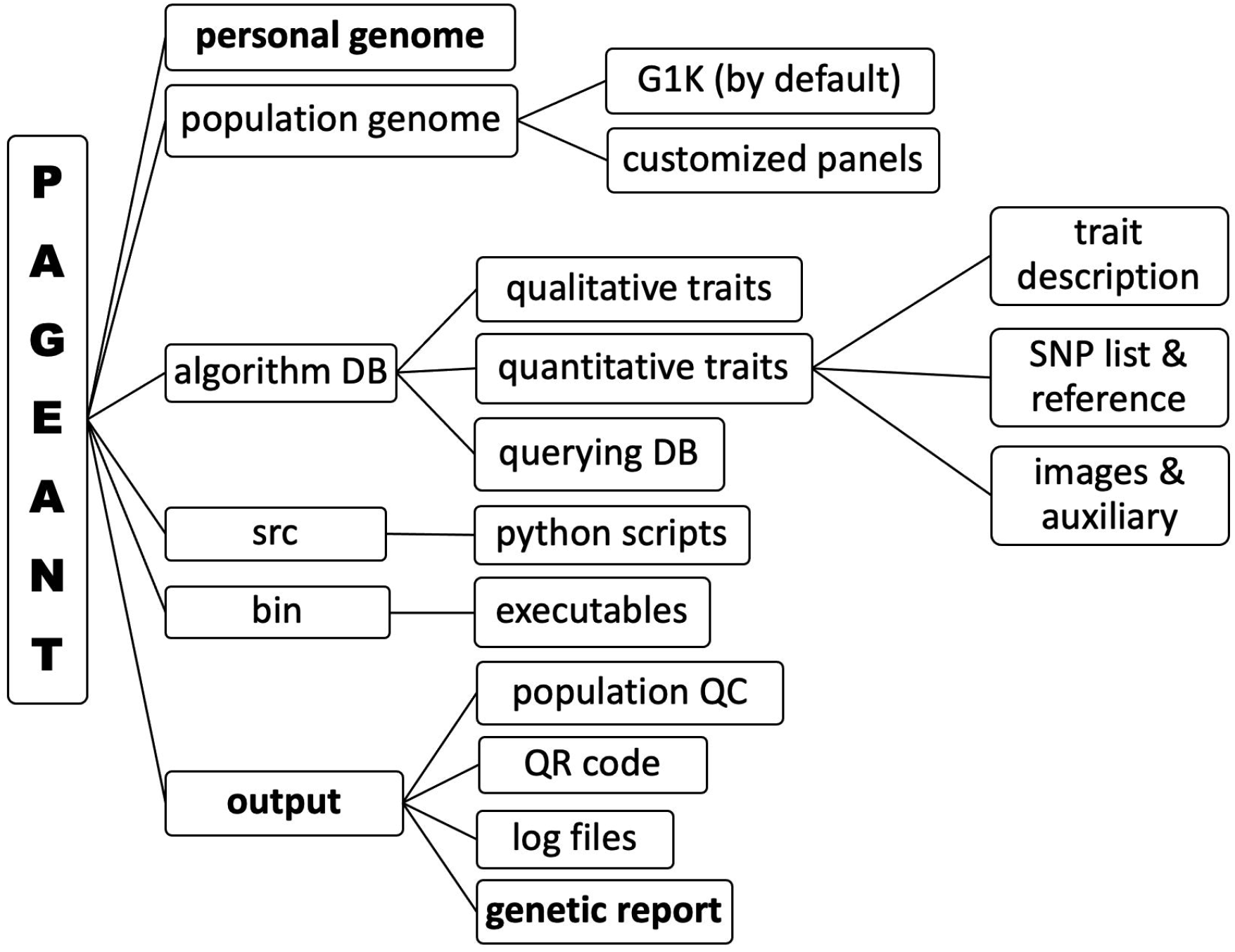
File structures outline when the software is locally installed. Advanced users could also follow this structure to customize the genetic report.

**Figure 3.**
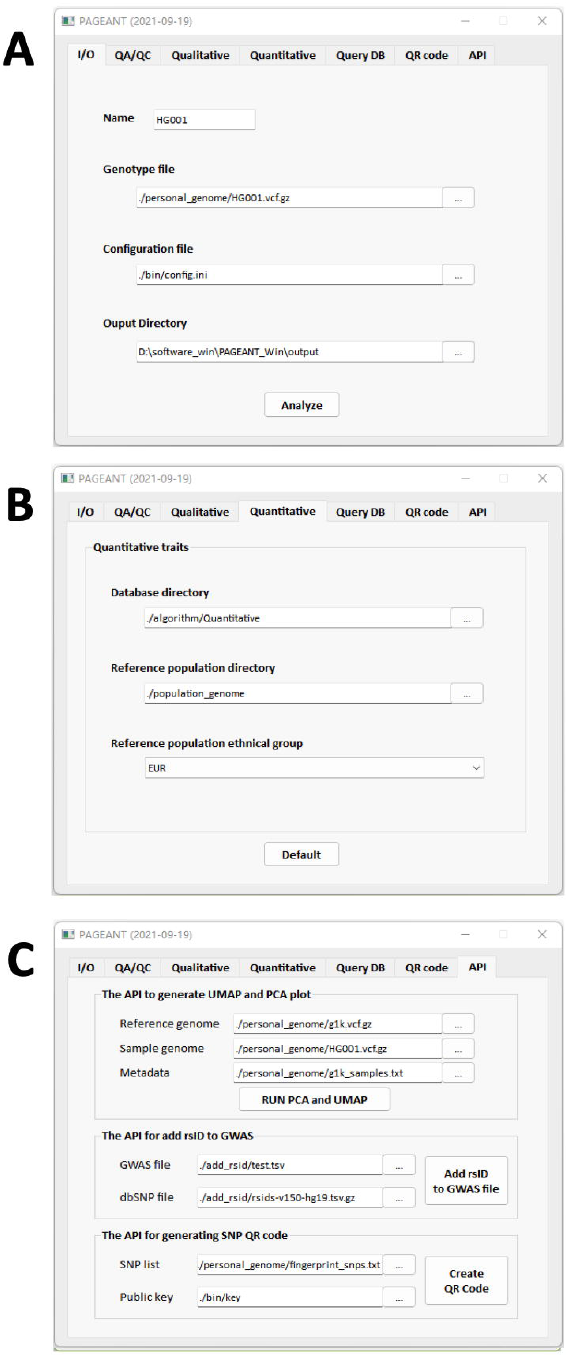
GUI interface of PAGEANT. (A) Main page “I/O”; (B) “Quantitative” page, where the user should select the appropriate ethnical group to obtain reliable PRS; (C) “APIs” page.

### 1^st^ Q: Quality assurance and quality control (QA/QC) report of genetic data

By leveraging the PLINK implementation in the PAGEANT software architecture, a basic QC is first performed, including genetically determined sex and the overall missing rate of the user’s genotype data, as basic quality assurance. As illustrated in **Figure 4**, the sample QC report also includes per chromosome distribution of detected variants along with ancestral positioning of personal genome based on Principal Component Analysis {PCA} and Uniform Manifold Approximation and Projection {UMAP} using the provided population reference (e.g., G1K). A dedicated API is implemented, which could be run easily since the required accessory libraries are already included once PAGEANT is installed (**Figure 3C**).

**Figure 4.**
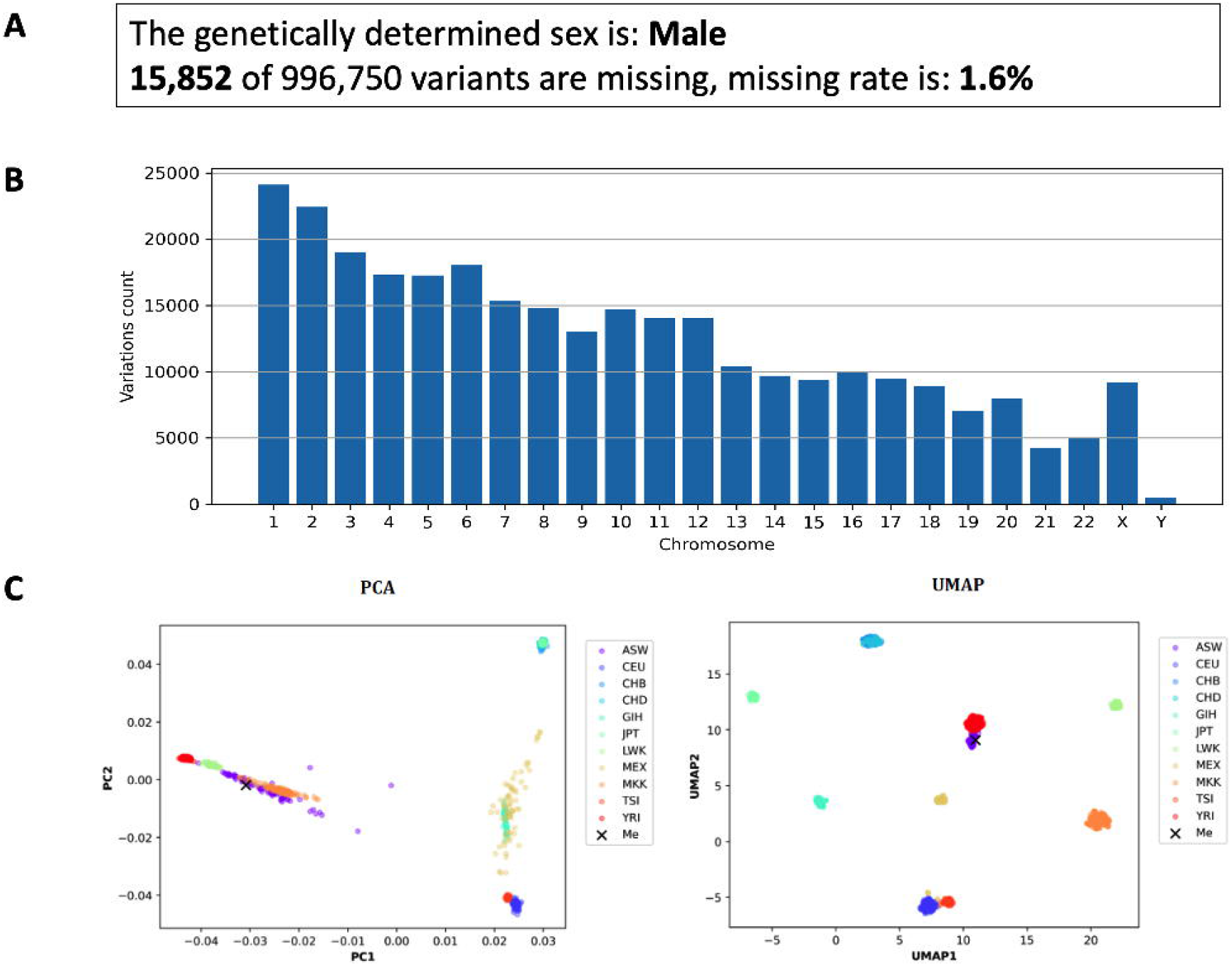
Sample QC report of PAGEANT. (A) Genetically determined sex and missingness rate; (B) Per chromosome count of detected variants; (C) Ancestral positioning of the personal genome based on Principal Component Analysis {PCA} and Uniform Manifold Approximation and Projection {UMAP} using the provided population reference (e.g., G1K).

### 2^nd^ Q: Qualitative assessment for genetic characteristics of absolute or relatively high certainty

This part of the genetic report is intended to present a limited list of genetic characteristics that could be reliably derived and that are of great interest to users. For example, one would want to know his ABO blood type, whether a sprinter or a muscular type person. By default, PAGEANT provides genetic reporting for a list of traits that the authors deemed eligible based on a literature review (**Figure 5A**).

**Figure 5.**
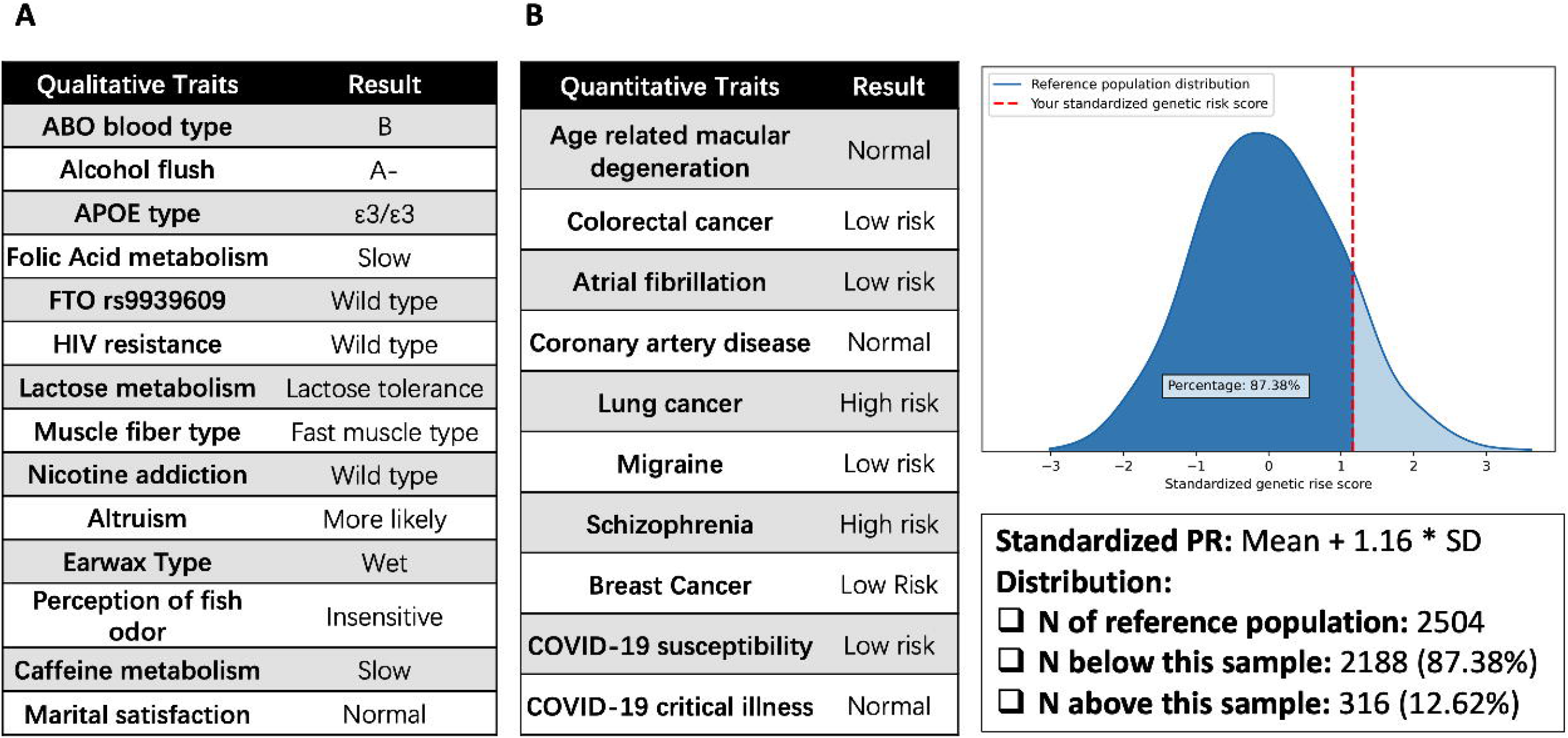
Qualitative and quantitative trait report output. PAGEANT provides (A) genetic reporting for a list of qualitative traits that the authors deemed eligible based on a literature review, and (B) quantitative trait scoring for polygenic traits based on most up to date GWAS literature.

### 3^rd^ Q: Quantitative trait scoring for polygenic traits based on most up to date GWAS literature

When PAGEANT is first launched, the 2,504 samples from G1K will have their traits processed first, so that the input individual genomic data has a population reference to measure each trait’s relative position among the entire G1K cohort (**Figure 5B**). One big advantage and innovation of this PAGEANT module is that advanced users could select their preferred GWAS file to calculate PRS. This should be more powerful than those provided by commercial vendors, because their PRS calculation is usually based on a few SNPs and users will not be able to customize it. Raw GWAS files usually come with millions of rows. Besides pruning, the biggest obstacle to adopt a GWAS like this into PAGEANT is that the SNP identifier is different between personal genome and population reference genomes. For example, the reference genome uses rsID as identifier, while many publicly released GWAS files use CHR:POS:REF:ALT format as identifier. Usually this takes an experienced bioinformatician to obtain the SNP identifier format aligned, especially for a GWAS with millions of records. This important function is implemented as an easy-to-use API (**Figure 3C**).

### 4^th^ Q: Query of 3^rd^ party variants databases of interest

As of February 29, 2020, the US National Institutes of Health (NIH) National Center for Biotechnology Information (NCBI) Genetic Testing Registry contained 64,860 genetic tests for 12,268 conditions and 18,686 genes from 560 laboratories (www.ncbi.nlm.nih.gov/gtr). The US Food and Drug Administration (FDA) had 404 entries for pharmacogenomic biomarkers used in drug labeling (www.fda.gov/drugs/science-and-research-drugs/table-pharmacogenomic-biomarkers-drug-labeling) and published a list of DTC tests with marketing authorization (https://www.fda.gov/medical-devices/vitro-diagnostics/direct-consumer-tests). The default version of PAGEANT allows users to query their genotype data for variants listed in these existing databases thus quickly identify genetic variants of interest (**Figure 6**).

**Figure 6.**
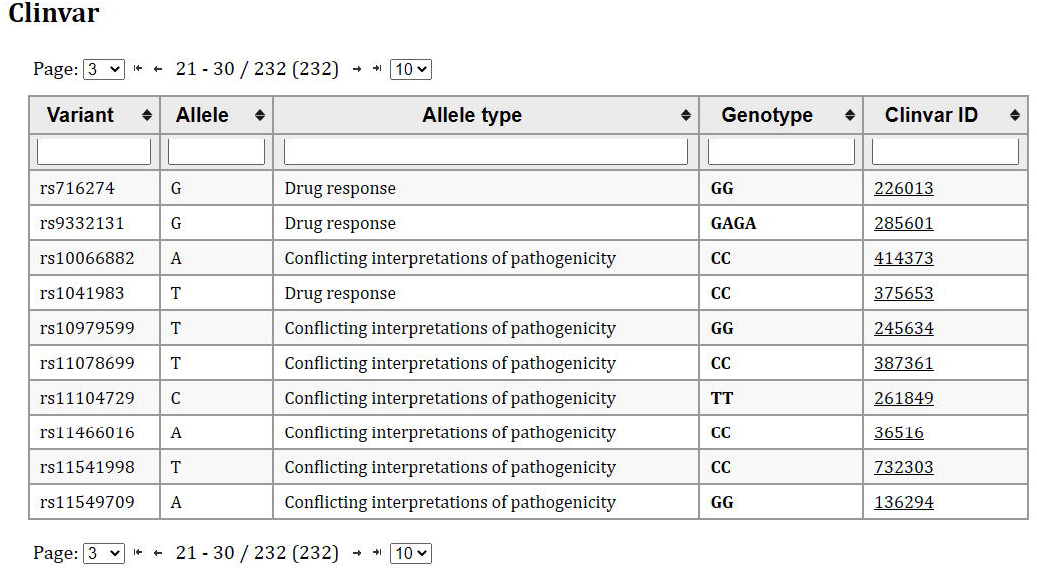
Query of 3rd party variants databases of interest (PharmKB, ClinVAR).

### 5^th^ Q: Quick Response (QR) code generation for specified genetic variants

For PAGEANT to extract and transmit a limited amount of genetic data in a convenient approach, we use QR code to code/decode. There are two QR codes involved: the first one is “public QR code”, encoding the list of SNPs (for example, 80 fingerprinting SNPs); the second one is “private QR code”, encoding the actual genomic data of a person for those SNPs coded in the “public” QR code. To further make this process secure, we implemented the Data Encryption Standard (DES) algorithm for data encryption/decryption on top of coding/decoding. There are two keys involved: the first one is “public key”, which is coded together with the list of SNPs in the public QR code; the second one is “private key”, which is hold only by the person who are authorized to access the limited person genome data. When a user scans the “public QR code”, PAGEANT decodes and decrypts it through DES algorithm, extract the genotype data for the list of SNPs, and then encrypts the extracted genotype data to generate his/her “private QR code”. This QR code could then be scanned and decrypted only by whoever holds the “private key”. **Figure 7** presents two QR codes for the scenario described above: a “public QR code” that encodes the “public key” together with a list of SNPs (on the left), a “private QR code” that encodes the user’s genetic data (on the right). A related API for generating a “public QR code” that encodes a “public key” is also available (**Figure 3C**).

**Figure 7.**
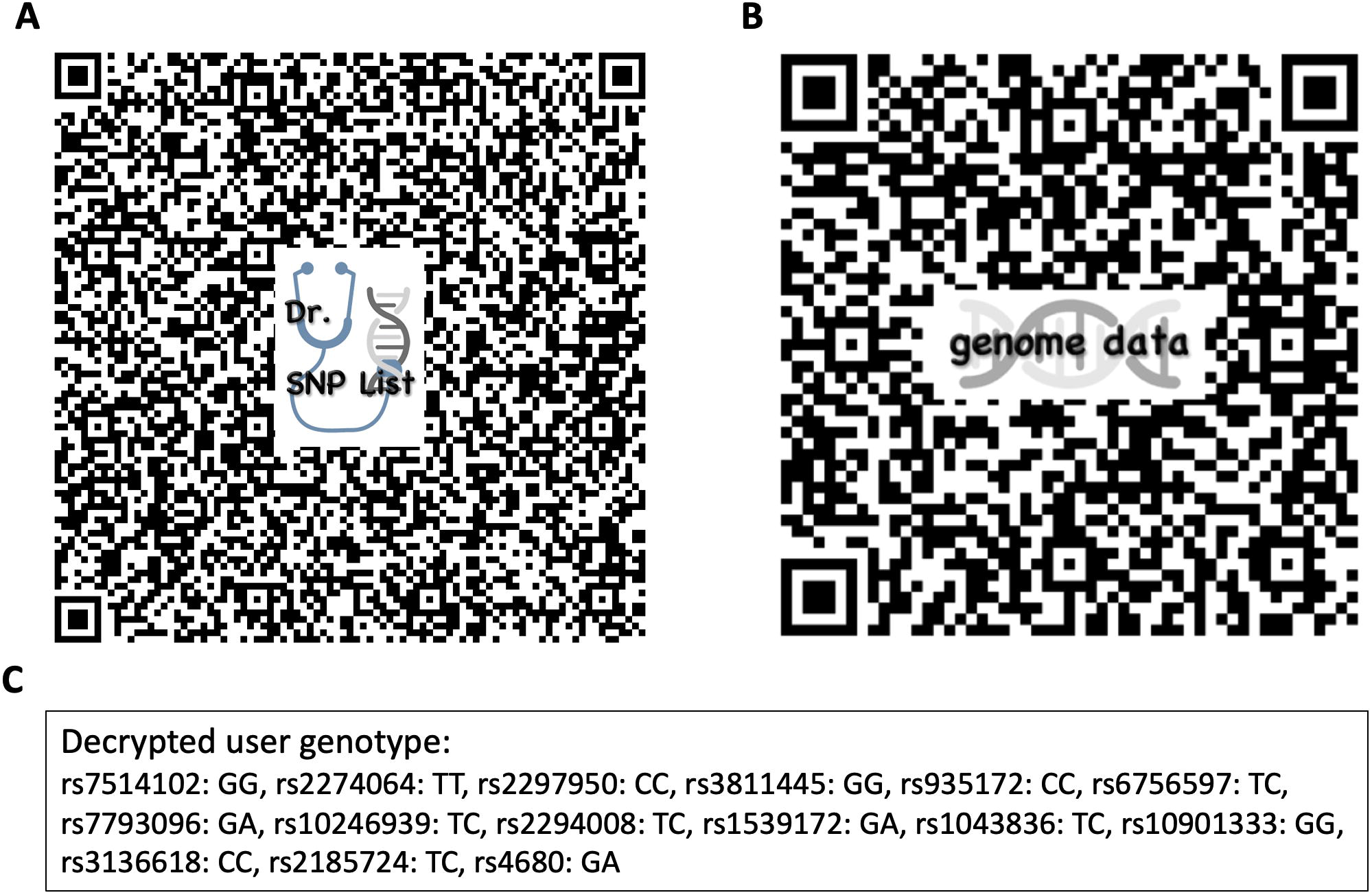
Quick Response (QR) code generation for specified genetic variants. (A) QR code for public key and SNP list; (B) QR code for user’s genotype data; (C) Decrypted user genotype.

## Discussion

Currently, DTC genetic testing is typically provided by commercial companies such as Ancestry.com (https://www.ancestry.com/), 23andMe (https://www.23andme.com) and MyHeritage (https://www.myheritage.it/dna). These vendors offer panels which include not only PRS but also carrier status and ancestry records. All these panels are generated starting from DNA taken from a saliva or blood sample then subjected to genotyping on genome-wide chips of up to 1 million variants. Up to the end of 2018, it has been estimated that 26 million people had used those online DTC companies [**4**]. Although several consumers are initially interested in ancestry research, they may later opt to use their raw genotype data to explore in third-party interpretation programs to analyze their genetic data for health purposes [**32**]. The information about life itself is undoubtedly much more abundant now and more valuable than “Google-able” information such as texts and images. However, the interpretation of genetic testing should not mainly rely on those driven by commercial interest and unscientific or inadequate evidences. It is the academic field that is making discovery for genetic mystery of human traits, and we strive to provide an academic version of tool that facilitates the translation of such science into personal access and knowledge. The vision of 6P medicine (participatory, predictive, preventive, personalized, precision, and policy) will forge a big step forward, when the DTC field now focus on getting more and more consumers to participate.

As a proof of concept and demonstration of technological feasibility, we developed PAGEANT (Personal Access to Genome & Analysis of Natural Traits), a DTC and DIY style of genetic reporting tool. PAGEANT is free to use and open for customization. It does not store users’ genotype data or mandate the way how the PRS is calculated. Although we provide a default prediction model for a few common traits as a reference by utilizing published results from GWAS Catalog, we also allow users to customize or even completely design their own model.

We also explored how to utilize the widely adopted QR code to securely transmit a small amount of personal genetic data. Previously, researchers developed Medicine Safety Code service to enable physicians and patients to represent pharmacogenomic data in QR code at the point-of-care [**33**]. The approach implemented in PAGEANT differs for two aspects: 1) PAGEANT allows extracting genotype in real-time, based on physician’ list of SNPs; 2) PAGEANT implements encryption/decryption besides coding/decoding, which is important for private patient genetic data. A patient has full control over all his/her private genomic data, only giving necessary genotype to the physician. And the physician also has full control on the medical interpretation of genetics. For example, if the patient has a pathogenic mutation for cancer, the physician may decide to not show it to the patient. As already stated in the introduction, it is not within the aims of the present paper to discuss about the pros and cons of DTC genetic testing or the ethical implications when getting genetically tested. A quite large literature is available on this matter [**14-18,27,31**].

Overall, PAGEANT represents a new, publicly available tool for third party genetic interpretation, that is totally transparent in its functionalities, so that the source code provided can be used for direct customization by the user and to expand the general knowledge about the “secrets” behind DTC genetic testing results interpretation. The output genetic report enabled displaying the log information of running the program so that users can quickly make sense of the underlying process and spot potential bugs. We want to highlight the great potential of PAGEANT also in the didactive context, by helping in training, preparing, and informing the next generation of scientists and clinically trained professionals that will face the ongoing race in personalized medicine businesses.

## Data Availability

PAGEANT code is available at https://github.com/jielab/pageant

## Data Availability

PAGEANT code is available at https://github.com/jielab/pageant

## Acknowledgements

This research was conducted using publicly released genome data from the 1000 genomes project (https://www.internationalgenome.org/). We acknowledge the participants of the 1000 Genomes Project to make their genome data available for the research community. We also utilized publicly released GWAS summary statistics from the COVID-19 host genetics initiative (https://www.covid19hg.org). Our hearts are with those who are affected by the COVID-19 pandemic.

